# The prevalence of SARS-CoV-2 antibodies within the community of a private tertiary university in the Philippines: a serial cross sectional study

**DOI:** 10.1101/2022.04.25.22274280

**Authors:** Lourdes Bernadette Sumpaico-Tanchanco, Jenica Sy, Angel Belle C. Dy, Myla Levantino, Arianna Maever L Amit, John Wong, Kirsten Angeles, John Paul Vergara

**Author notes:** Corresponding author (LBST).

## Abstract

The antibody testing for severe acute respiratory syndrome coronavirus 2 (SARS-CoV-2) was used to detect the presence of antibodies in a private university setting. This serial cross-sectional study determined the seroprevalence of SARS-CoV-2 antibodies using qualitative and quantitative tests. Between June 2021 to December 2021, samples from 1,318 participants were tested, showing 47.80% of the study population yielding IgG antibodies to SARS-CoV-2 virus. A general increase in seroprevalence was observed from June to December 2021. However, a decreasing trend in IgG reactivity was found in vaccinated individuals over time. IgG antibody formation was observed across all brands of vaccines.

## Background

The coronavirus disease (COVID-19) is caused by the severe acute respiratory syndrome coronavirus 2 (SARS-CoV-2) virus which has infected millions of people. A comprehensive understanding of the virus and how it spreads is crucial in anticipating the epidemiology of COVID-19 to make informed healthcare and financial decisions (1,2).

Serology, or antibody, testing determines the presence of antibodies against SARS-CoV2 in an individual’s blood sample (3). Seroprevalence surveys estimate the percentage of individuals in a population who have antibodies against the virus at a large scale. Antibodies are produced in the blood by the body’s immune system when there are harmful substances it needs to protect, in this case the COVID-19 infection. Serological testing is ideal in approximating cumulative prevalence because antibodies against SARS-CoV2 persist for a longer amount of time, after infection (4).

Population-based seroprevalence studies have been conducted in hospital settings and community settings in countries across the world, such as China (4), Switzerland (5), Iran (6), Hong Kong (7), US (8), and Brazil (9), among others. Studies have been conducted cross sectionally, across different households and age groups, using immunofluorescence assays for anti-SARS-CoV2 IgG and IgM antibodies. In England, a large-scale seroprevalence study, named the Real-time Assessment of Community Transmission (REACT) study is currently underway. The REACT program is being conducted at the national seroprevalence level to monitor the persistence of SARS-CoV2 antibodies through at-home antigen swab tests and finger-prick antibody tests. The first phase of the study, which utilized the antigen tests, involved more than 150,000 individuals, and the second phase, which utilizes the finger prick antibody tests, is currently ongoing (1). A multi-country study involving the seroprevalence of COVID-19 antibodies, called Solidarity II, is also being conducted by the World Health Organization (WHO) in order to fill in the knowledge gaps of the global epidemiology of COVID-19.

Antibody testing is instrumental in generating epidemiological information necessary for the control of infectious diseases, including COVID-19. The surveillance of COVID-19 is necessary in order to determine disease burden in the population and to detect vulnerable groups who are at high risk. Determining the prevalence of COVID-19 is important in understanding the widespread outbreak of the disease, transmission of the virus, and the immunity status of the population. In order to implement proper strategies to eradicate COVID-19, it is crucial to identify the number of people previously and currently infected with the virus, and who are most at risk.

Widespread COVID-19 testing can help the country determine the true state of infection of the community and identify hotspots and high-risk populations. However, large-scale molecular testing, such as RT-PCR detection, is not easily achievable due to limitations in testing kits and expenses. At the time of this study, there were initially 219 laboratories that were accredited by the Department of Health (DOH) to conduct COVID-19 RT-PCR testing, which increased to 280 laboratories in October 2021 (10). There was an average testing of about 50,000 to 58,000 daily, and positivity rates ranged from 10.6% to 28.1% from June to October 2021.

Antibody testing is a reliable diagnostic alternative that can be used (11). It is a preferable option in testing at a large-scale, especially if the tests used exhibit quality performance. It involves a simple lateral flow test that only requires a minute amount of blood with no lab processing. This assay detects antibody responses that can be used to diagnose acute infection. It can be used to detect binding antibodies and identify recovered cases through remaining IgG antibodies (12). SARS-CoV2 infected patients mostly exhibit an antibody response 10-15 days after infection, and there is a sequential or simultaneous seroconversion for immunoglobulin G (IgG) and immunoglobulin M (IgM) (13). Quantitative antibody tests may determine antibody titers, enable longitudinal monitoring of antibody levels in patients, and potentially monitor antibody response to vaccines.

The primary objective of this study is to determine the seroprevalence of antibody titers in employees in an tertiary academic university across a 6 month period. The secondary objective is to determine the association between sociodemographic characteristics, familial history, vaccine brand, COVID-19 testing positivity, and serological status. Finally, the study aims to correlate qualitative and quantitative antibody testing. No previous COVID-19 antibody seroprevalence study in a university community has been conducted in the Philippines.

## Methods

### Study design

A serial cross-sectional study was conducted from June to December 2021, when the vaccination roll-out started in the Philippines. Participants were recruited for a rapid antibody test once per month. In the third month, a qualitative antibody test was performed along with the qualitative rapid antibody test.

### Participant recruitment

Participants of the study included faculty, administrators, professionals, staff, maintenance, security guards, affiliates, in-campus residents, and students of Ateneo de Manila University (AdMU). They were recruited through online avenues of communication through official school channels. All participants accomplished a study questionnaire consisting of questions related to each of their demographics, medical history, COVID-19 vaccination status, and COVID-19 precautionary behaviors.

The computed sample size was at least 400 to ensure that the 95% CI will be an interval with length less than 0.10 (point estimate ±< 0.05 margin of error) (14,15). However, a total of 500 participants was targeted for recruitment to account for potential dropouts.

The study was reviewed and approved by the Institutional Review Board (IRB) from the School of Medicine and Public Health Panel of the AdMU Research Ethics Committee. Informed consent was secured prior to inclusion to the study through the digital data collection tool used in the study.

### Data collection and monitoring instrument

BluEHR (Tantum Quantum Headquarters, Inc), a cloud-based electronic medical record (EMR) was used as the main data collection tool for this project.

### Sample collection and antibody testing

A medical professional (medical technologist or physician) collected blood samples from the participants. The Roche SARS-CoV-2 Rapid Antibody Kit was used for the qualitative antibody testing. It has a specificity of 100% and sensitivity of 92%. Twenty (20) μl of blood was obtained through finger pricking. The collected blood was added to the test device, followed by 90 μl (approximately 3 drops) of buffer. The test was incubated for 10-15 mins before interpretation. Participants with reactive IgM results were referred to the university physician as it was beyond the scope of this study to do confirmatory rt-PCR testing.

The Roche Quantitative Anti-SARS-COV-2 S Antibody testing kit was used for quantitative antibody testing in the last month of collection. Three to 4 ml of blood were obtained from each participant through venipuncture. The samples were stored in a vacutainer tube and sent to an outsourced laboratory for testing while stored at 2-4°C. Results were interpreted as follows: values <0.8 U/mL are considered non-reactive, and values ≥0.8 U/mL are considered reactive. The outsourced laboratory is licensed for operation by the Department of Health and is also registered at the National Privacy Commission.

### Data analysis

Seroprevalence was defined as the proportion of participants with a reactive IgG result in the qualitative rapid antibody test (16). The data was analyzed using SPSS software (version 28.0.1.0) and Google Sheets.

Univariate analysis was conducted to provide the descriptive statistics of the study population. Frequency distributions were made for categorical variables, and the mean, standard deviations, median, and quartiles were obtained for continuous variables (i.e. age and quantitative count). Bivariate analyses were performed using chi-square tests, independent t-tests, and a one-way analysis of variance (ANOVA). This was used to look at the relationship between categorical variables, in addition to the logistic regression. Binomial logistic regression modeling was also performed as a multivariate analysis, to obtain odds ratios and determine the relationship between the dependent variable with one or more independent variables. Results were considered statistically significant if p-values were less than 0.05.

The following nonbinary variables were collapsed into dichotomous variables to fulfill the assumptions for the chi-square tests (to compare the two groups with respect to the outcome variables) and were also used for the multivariate tests:

1. Age: collapsed into ages <60; and ≥60
2. Region of residence: collapsed into living into NCR; and outside NCR
3. Highest educational attainment categories: collapsed into having attained college/postgrad education; and others (i.e. elementary, high school, vocational, no formal education)
4. Work arrangement: collapsed into working remotely; and onsite/hybrid
5. Having tested positive to COVID-19: collapsed into having tested positive (regardless of symptom severity); and not
6. Having at least one individual in the household testing positive with COVID-19: collapsed into a household individual having had tested positive (regardless of symptom severity); and not

Other variables were already dichotomous (i.e. sex, comorbidities). One-way ANOVA was used to analyze a relationship between the quantitative count and vaccine brands. Tukey’s HSD was used as a post hoc test.

In order to compare the results of the qualitative and quantitative antibody tests, a receiver operating characteristic (ROC) analysis was performed (17,18). The most plausible U/ml cutoff for the qualitative test kit was estimated by considering each measurement as a candidate cutoff point and computing the resulting accuracies. In this specific analysis, accuracy is defined as the percentage of correctly classified data points over N=291, where “correctly classified” is determined by using the results obtained from the qualitative test as ground truth.

## Results

A total of 1,318 participants were recruited to participate in the study. The number of participants per month is summarized in Table 1.

**Table 1.** Number of participants per round of data collection.

| Round | N |
| --- | --- |
| June | 459 |
| July | 158 |
| September | 198 |
| October | 457 |
| December | 46 |

Among the participants, 50.8% were males and 49.2% were females. The study population ages ranged from 20 to 87, with a median age of 38 years (SD±13.1). Majority of them were college graduates or with postgraduate degrees (87.5%). Work arrangements of the participants also varied in that 33.1% worked remotely, while 55.5% had onsite work. Almost half (49.5%) of participants reported having at least one comorbidity, with hypertension being the most common (Table 2).

**Table 2.** Sociodemographic characteristics of the study population (N=1,318).

| Demographics | N (%) |
| --- | --- |
| Sex |  |
| Female | 648 (49.2) |
| Male | 670 (50.8) |
| Age group |  |
| 18-59 | 1244 (94.4) |
| ≥60 | 74 (5.6) |
| Role in the University |  |
| Administration/ Professional | 179 (13.6) |
| Faculty | 347 (26.3) |
| In-campus Residents | 42 (3.2) |
| Maintenance/ Guards/ Subconcessional/<br>Outsourced/ Diars/ Affiliates | 211 (16) |
| Staff | 267 (20.3) |
| Student | 272 (20.6) |
| Educational attainment |  |
| College/ Postgraduate program | 1,153 (87.5) |
| Primary/ Secondary education/ Others | 146 (11.1) |
| Missing | 19 (1.4) |
| Work arrangement |  |
| Remote | 436 (33.1) |
| On-site | 731 (55.5) |
| Missing | 151 (11.5) |
| Area of Residence |  |
| NCR | 1,074 (81.5) |
| Outside NCR | 244 (18.5) |
| Comorbidities |  |
| None | 516 (39.2) |
| With at least 1 comorbidity | 652 (49.5) |
| Diabetes | 104 (7.9) |
| Hypertension | 260 (19.7) |
| Obesity | 137 (10.4) |
| COPD | 1 (0.1) |
| CVD | 4 (0.3) |
| Asthma | 84 (6.4) |
| Missing | 150 (11.4) |
| Smoking status |  |
| Smoker | 97 (7.4) |
| Non-smoker | 1,071 (81.3) |
| Missing | 150 (11.4) |

During the antibody testing, only 37 (2.81%) had reactive IgM serology results. Six hundred thirty (47.80%) participants had reactive IgG serology results. Of those who received a reactive IgG result, 486 (77.14%) never tested positive for COVID-19 through RT-PCR. In our study population, only 38 (2.9%) participants previously tested positive for COVID-19 through RT-PCR, and 1,243 (94.3%) have not tested positive. The seroprevalence of the study population ranged from 28.76% to 65.15%. A general increase in trend can be seen in the seroprevalence of participants over time.

From the participants with reactive IgG results, those who have had COVID-19 were excluded to determine the seropositivity rates caused by vaccination. Participants who received their vaccine more than 6 months prior to testing were also excluded due to the small sample size (n=17). The seroprevalence of vaccinated members of the university community during the study period ranged from 29%-69.40%, across 6 months, shown in Fig 2. A general decrease in antibody levels was found over time. Among men, the seroprevalence was 18.89%, and among women, it was 18.74%.

**Fig 1.** Seroprevalence of the study population across batches.

**Fig 2.** Percent of vaccinated respondents with reactive IgG results across time.

Excluding participants who were previously infected, the total antibody titers ranged from 0.4 U/mL to >25,000 U/mL in the quantitative test (n=409). The mean was 3,099.56 U/mL across all brands, and the descriptives are further summarized in Table 3.

**Table 3.** Descriptive summary of the quantitative antibody test results according to vaccine brand.

| Vaccine Brand | N | Mean | Std. Deviation | 95% CI for Mean |  | Minimum | Maximum |
| --- | --- | --- | --- | --- | --- | --- | --- |
|  |  |  |  | Lower Bound | Upper Bound |  |  |
| Astrazeneca | 88 | 2652.07 | 5308.56 | 1527.29 | 3776.85 | 9.99 | 25000.00 |
| Johnson and Johnson | 4 | 2922.32 | 4439.41 | -4141.76 | 9986.41 | 230.00 | 9544.00 |
| Moderna | 29 | 7916.83 | 8177.15 | 4806.40 | 11027.25 | 388.00 | 25000.00 |
| Pfizer Biontech | 28 | 3292.21 | 5314.19 | 1231.58 | 5352.84 | 185.00 | 25000.00 |
| CoronaVac | 256 | 1591.17 | 4837.28 | 995.78 | 2186.55 | 0.40 | 25000.00 |
| Sputnik | 4 | 931.37 | 450.38 | 214.72 | 1648.03 | 294.00 | 1284.00 |
| Total | 409 | 2390.97 | 5468.42 | 1859.42 | 2922.51 | 0.40 | 25000.00 |

The results of the qualitative antibody test were correlated with the quantitative test in order to identify a cutoff value at which participants received a reactive IgG result. Fig 3 displays the resulting accuracies against the candidate cutoff points and it reveals that a threshold of 169.5 U/ml yielded maximum accuracy (89.35%). This means that a positive result from the qualitative test occurs when at least 169.5 U/ml is detected from the specimen (note that the candidate value before this cutoff is 152, so alternatively, the condition can be stated as > 152).

**Fig 3.** Percentage of accuracy for the quantitative antibody test cut off value.

The cutoff value was also verified using ROC analysis and the Youden index (15,16). This yielded the same optimal cutoff point, as shown in Fig 4. It is important to note that this value is an indication of the cutoff value at which both the qualitative and quantitative tests give reactive results. Therefore, it does not determine the accuracy of the tests used and is not conclusive of a definite level of protection against infection.

**Fig 4.** ROC analysis and Youden index of the cutoff value.

Table 4 shows the contingency table comparing the qualitative results against the quantitative results using the 169.5 U/ml threshold.

**Table 4.** Contingency table comparing the qualitative and quantitative results.

| Quantitative Count |  |  |  |
| --- | --- | --- | --- |
| (U/ml) | Positive | Negative | Total |
| $\geq 169.5$ | 167 | 11 | 178 |
| $< 169.5 (\leq 152)$ | 20 | 93 | 113 |
| Total | 187 | 104 | 291 |

Participants with obesity or those who are smokers were found to be 1.7x and 1.8x more likely to have less reactive IgG serology results, respectively. Apart from this, there were no significant associations with other risk factors and IgG reactivity.

Since the first sample collection in June until December 2021, 187 (14.2%) participants received their first dose, 878 (66.6%) were fully vaccinated, and 253 (19.2%) were unvaccinated. Most of the participants (48%) received CoronaVac as their vaccine followed by AstraZeneca at 15% of participants. Johnson and Johnson, and Sputnik were excluded from this analysis due to the small number of individuals who received them. It was observed that the Pfizer vaccine produced the highest amount of antibodies, followed by Moderna, Astra Zeneca and CoronaVac in decreasing order. Those who tested positive on the IgG test are 58.1x more likely to have received Pfizer vaccine, and so on. It is important to note that those who were vaccinated, regardless of vaccine brand, were more likely to have reactive IgG serology values (Table 6).

**Table 5.** Adjusted odds ratios of risk factors affecting IgG serology among study participants.

| Risk Factor | p | a OR (95% CI) |
| --- | --- | --- |
| Sex | 0.075 | 1.314 (0.972-1.775) |
| Ages <60 and ≥60 | 0.689 | 0.997 (0.984-1.011) |
| Region of Residence | 0.705 | 1.079 (0.728-1.601) |
| Highest Educational Attainment Categories | 0.292 | 0.769 (0.472-1.254) |
| Work Arrangement | 0.122 | 0.775 (0.562-1.070) |
| Comorbidities |  |  |
| With at least 1 comorbidity | 0.922 | 1.023 (0.649-1.613) |
| Diabetes | 0.541 | 0.850 (0.505-1.430) |
| Hypertension | 0.136 | 0.705 (0.446-1.116) |
| Obesity | 0.047* | 0.603 (0.366-0.994) |
| Asthma | 0.411 | 0.775 (0.421-1.424) |
| Smokers | 0.021* | 0.543 (0.323-0.913) |
| Household Testing Positive | 0.614 | 1.231 (0.549-2.761) |
| At Least 1 Household Individual Vaccinated | 0.066 | 1.492 (0.974-2.287) |
| Vulnerable Individuals in Household | 0.462 | 0.892 (0.657-1.210) |
| Frontliners In Household | 0.096 | 1.306 (0.953-1.790) |

\*Statistically significant data (p < 0.05).

**Table 6.**
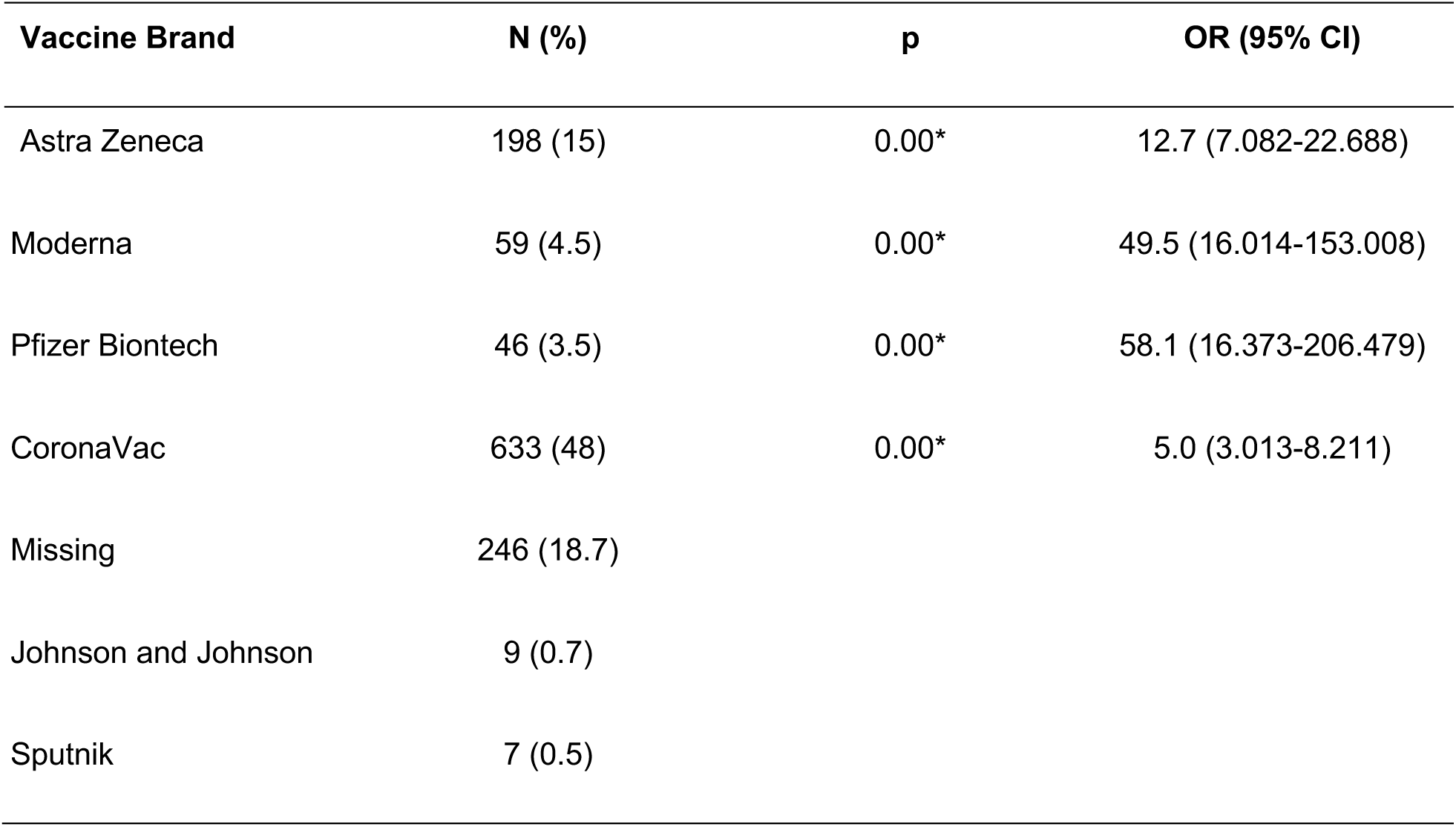
IgG reactivity with vaccine brand based on the qualitative antibody test.

## Discussion

This study was conducted in a university community with the majority of participants holding baccalaureate and post-graduate degrees. During this time only half of the workforce was entering the campus for onsite work. Vaccination roll-out just started when the study was initiated.

Seroprevalence is an estimate of people who have developed antibodies against a pathogen. In a review by Post and colleagues, it was found that seroconversion of IgM antibodies occurred any time from 4 to 14 days after infection and declined 6 weeks after disease onset (19). On the other hand, IgG antibodies were detected from 12-15 days after infection, and declined after 8 weeks.

We observed a seroprevalence rate that ranged from 28.76% to 65.15% among members of the community, from June to December 2021. There was no significant difference in seroprevalence by gender and age in the study population. Seropositivity in the analysis was mainly due to the antibody response developed due to vaccination. These values are consistent with results published regarding COVID-19 vaccine development. IgG titers against the SARS-CoV2 S protein, spike subunit, and receptor binding domain have previously been measured in individuals with mRNA vaccines, developed by Pfizer Biontech or Moderna (20–23). It was found that more than 97% of individuals developed IgG titers after their first dose, and 100% of them developed antibodies after their second dose. Studies conducted on the vaccines with the inactivated virus yielded similar results. A study on 1,247 healthcare workers in Brazil showed that there was an increase in seropositivity rates after the first dose (88%), and an even higher seropositivity rate after the second dose (99%) (24).

Although vaccination rates were high across the study population, a decreasing trend in the seroprevalence was found over time after vaccination. After the development of an antibody response to the S protein of the virus, a decline in antibody levels has been observed after 3 months and after 6 months of the second dose (20,24,25). This may explain the decline in seroprevalence found in our results over time. There was a decline in seropositivity rates from vaccination. This decrease of antibodies over time supports the need for booster shots that was recommended by the Department of Health. This also supports the continuous use of health precautions like use of masks, frequent handwashing and social distancing.

Among the risk factors included in the study, only obesity and smoking status were significantly associated with IgG serology. Similar to this, a seroprevalence study in Switzerland (5) and Russia (26) showed that current smokers were less likely to have reactive serology results compared to non-smokers, though these studies only focused on seropositivity after infection. Chronic health conditions were also not significantly associated with serology results in the Switzerland population (5). However, contrary to our results, this study also reported that obese women had higher odds of receiving reactive serology results compared to non-obese women, but there were no differences observed in male participants. A cross-sectional seroprevalence survey in Abu Dhabi also resulted in similar results, with smokers showing lower seroprevalence (27). BMI categories and comorbidities did not show any significant associations with seropositivity.

This study was limited to the measurement of antibodies against SARS-CoV2 in a university population. It was important to note that antibody seroprevalence varied depending on the location, target participants, and timing in which the testing was conducted during the pandemic (28). The seroprevalence obtained in this study was only reflective of the status of the university community at the time of testing. Antibody testing was not used to diagnose infection in individuals. At the moment, there was no consensus on the best approach used to evaluate COVID-19 infection status in relation to a university community (29,30). Donneau and colleagues recommended the use of a saliva RT-PCR test and antibody test in conjunction with self-reporting of symptoms in order to determine the prevalence of COVID-19 in the population and characterize seroconversion and seroreversion after vaccination and infection (31). A case study on Cornell University used 3 mechanisms to limit transmission: testing, contact tracing, and symptomatic self-reporting. The study recommended that universities test students at least once per week, regardless of vaccination rates and status (32). Similar recommendations were provided in modeling studies conducted at Duke and Harvard Universities. In addition, other preventive measures, such as campus-based screening, quarantine protocols, hygienic measures, and cafeteria and dormitory regulation, were also necessary for the safe reopening of universities (33). Local universities may continue to monitor COVID-19 antibody status as a measure of vaccination status for an epidemiological situational analysis. Antibody testing was reserved for research purposes only.

## Conclusions

This study conducted in a Filipino university showed a SARS-CoV2 antibody seroprevalence range of 28.76% to 65.15%. This study was conducted as the vaccination roll out in the Philippines started and involved individuals assuming different roles in the university. A decreasing trend of levels of antibody was observed in participants over the next 6 months after vaccination. IgG antibody formation was observed in all brands of vaccines. Among the brands, antibody reactivity was observed in Pfizer, Moderna, Astra Zeneca and CoronaVac, in decreasing order. The presence of comorbidities did not show an added possibility of having antibodies.

Because of the decreasing trend of antibody levels, it is recommended that vaccination status of individuals need to be monitored by university officials and guidelines for booster doses are followed. Since all the vaccines examined showed antibody positivity, these brands can be considered when available. Antibody testing is not recommended to monitor the status of current infection in the community. It can be used, however, for epidemiological surveillance purposes to assess herd immunity.

It is recommended that public health safety measures like wearing of masks, frequent hand washing, social distancing be continued. Contact tracing and monitoring especially in in-campus dormitories should be enforced. While vaccination is one of several methods that should be done to ensure safety against COVID-19, it is recommended that a surveillance system, such as waste water surveillance and air sampling, be implemented as environmental measures to monitor the spread of the virus. An effective science communication system should be enforced to keep the community informed of risk status in different activities in the university across different settings.

## Data Availability

Data cannot be shared publicly because these are being presented to university officials first. Data are available from the Ethics Committee (contact via) for researchers who meet the criteria for access to confidential data.

## Acknowledgments

This study was funded by the Ateneo de Manila University Office of the Vice Presidents of the Loyola Schools and the Professional Schools. We would like to acknowledge the university President and Vice Presidents and university Deans for their support of the research. We are grateful for the administrative and logistic support of the following people: Engr. Elias Pan, Ms. Joy Rodriguez-Salita, Mr. Darwin Enguerra, Ms. Kriselle Abcede, Ms. Maria Lourdes Almeda-Benito, Ms. Carolyn Cabardo and team, and the Campus Safety and Mobility Office. We are thankful to the Tantum Quantum Headquarters team for their help in setting up the BluEHR monitoring tool and the United Diagnostics Laboratory for their help in conducting the quantitative antibody tests. We are also thankful for all the university community members who participated in the study.

## References

1. Moshe M, Daunt A, Flower B, Simmons B, Brown JC, Frise R, et al. SARS-CoV-2 lateral flow assays for possible use in national covid-19 seroprevalence surveys (React 2): diagnostic accuracy study. BMJ. 2021 Mar 2;372:n423.

2. Krsak M, Johnson SC, Poeschla EM. COVID-19 Serosurveillance May Facilitate Return-to-Work Decisions. Am J Trop Med Hyg. 2020 Jun;102(6):1189–90.

3. CDC. atWhat COVID-19 Seroprevalence Surveys Can Tell Us [Internet]. Centers for Disease Control and Prevention. 2020 [cited 2021 Dec 6]. Available from: https://www.cdc.gov/coronavirus/2019-ncov/covid-data/seroprevalance-surveys-tell-us.html

4. Xu X, Sun J, Nie S, Li H, Kong Y, Liang M, et al. Seroprevalence of immunoglobulin M and G antibodies against SARS-CoV-2 in China. Nat Med. 2020 Aug;26(8):1193–5.

5. Stringhini S, Wisniak A, Piumatti G, Azman AS, Lauer SA, Baysson H, et al. Seroprevalence of anti-SARS-CoV-2 IgG antibodies in Geneva, Switzerland (SEROCoV-POP): a population-based study. Lancet Lond Engl. 2020 Aug 1;396(10247):313–9.

6. Shakiba M, Nazemipour M, Salari A, Mehrabian F, Nazari SSH, Rezvani SM, et al. Seroprevalence of SARS-CoV-2 in Guilan Province, Iran, April 2020. Emerg Infect Dis. 2021 Feb;27(2):636–8.

7. To KK-W, Cheng VC-C, Cai J-P, Chan K-H, Chen L-L, Wong L-H, et al. Seroprevalence of SARS-CoV-2 in Hong Kong and in residents evacuated from Hubei province, China: a multicohort study. Lancet Microbe. 2020 Jul;1(3):e111–8.

8. Havers FP, Reed C, Lim T, Montgomery JM, Klena JD, Hall AJ, et al. Seroprevalence of Antibodies to SARS-CoV-2 in 10 Sites in the United States, March 23-May 12, 2020. JAMA Intern Med. 2020 Jul 21;

9. Borges LP, Martins AF, de Melo MS, de Oliveira MGB, Neto JM de R, Dósea MB, et al. Seroprevalence of SARS-CoV-2 IgM and IgG antibodies in an asymptomatic population in Sergipe, Brazil. Rev Panam Salud Pública. 2020 Oct 6;44:e108.

10. COVID-19 Case Tracker | Department of Health website [Internet]. [cited 2022 Feb 22]. Available from: https://doh.gov.ph/covid-19/case-tracker

11. Peeling RW, Wedderburn CJ, Garcia PJ, Boeras D, Fongwen N, Nkengasong J, et al. Serology testing in the COVID-19 pandemic response. Lancet Infect Dis. 2020 Sep;20(9):e245–9.

12. Adams ER, Ainsworth M, Anand R, Andersson MI, Auckland K, Baillie JK, et al. Antibody testing for COVID-19: A report from the National COVID Scientific Advisory Panel. Wellcome Open Res. 2020;5:139.

13. Mahajan A, Manchikanti L. Value and Validity of Coronavirus Antibody Testing. Pain Physician. 2020 Aug;23(4S):S381–90.

14. Greenland S. Basic Methods for Sensitivity Analysis of Biases. Int J Epidemiol. 1996 Dec 1;25(6):1107–16.

15. Diggle PJ. Estimating Prevalence Using an Imperfect Test. Epidemiol Res Int. 2011 Oct 23;2011:e608719.

16. Tuells J, Egoavil CM, Pena Pardo MA, Montagud AC, Montagud E, Caballero P, et al. Seroprevalence Study and Cross-Sectional Survey on COVID-19 for a Plan to Reopen the University of Alicante (Spain). Int J Environ Res Public Health. 2021 Feb 16;18(4):1908.

17. Doi SAR. Using and Interpreting Diagnostic Tests with Quantitative Results. In: Doi Sar, Williams GM, editors. Methods of Clinical Epidemiology [Internet]. Berlin, Heidelberg: Springer; 2013 [cited 2022 Mar 12]. p. 67–78. (Springer Series on Epidemiology and Public Health). Available from: https://doi.org/10.1007/978-3-642-37131-8_6

18. Habibzadeh F, Habibzadeh F, Habibzadeh P, Yadollahie M. On determining the most appropriate test cut-off value: the case of tests with continuous results. Biochem Medica. 2016 Oct 15;26(3):297–307.

19. Post N, Eddy D, Huntley C, Schalkwyk MCI van, Shrotri M, Leeman D, et al. Antibody response to SARS-CoV-2 infection in humans: A systematic review. PLOS ONE. 2020 Dec 31;15(12):e0244126.

20. Naaber P, Tserel L, Kangro K, Sepp E, Jürjenson V, Adamson A, et al. Dynamics of antibody response to BNT162b2 vaccine after six months: a longitudinal prospective study. Lancet Reg Health -Eur. 2021 Nov 1;10:100208.

21. Stamatatos L, Czartoski J, Wan Y-H, Homad LJ, Rubin V, Glantz H, et al. mRNA vaccination boosts cross-variant neutralizing antibodies elicited by SARS-CoV-2 infection. Science [Internet]. 2021 Jun 25 [cited 2022 Feb 8]; Available from: https://www.science.org/doi/abs/10.1126/science.abg9175

22. Jalkanen P, Kolehmainen P, Häkkinen HK, Huttunen M, Tähtinen PA, Lundberg R, et al. COVID-19 mRNA vaccine induced antibody responses against three SARS-CoV-2 variants. Nat Commun. 2021 Dec;12(1):3991.

23. Wheeler SE, Shurin GV, Yost M, Anderson A, Pinto L, Wells A, et al. Differential Antibody Response to mRNA COVID-19 Vaccines in Healthy Subjects. Martinez MA, editor. Microbiol Spectr [Internet]. 2021 Sep 3 [cited 2022 Feb 9];9(1). Available from: https://journals.asm.org/doi/10.1128/Spectrum.00341-21

24. Fonseca MHG, de Souza T de FG, de Carvalho Araújo FM, de Andrade LOM. Dynamics of antibody response to CoronaVac vaccine. J Med Virol [Internet]. [cited 2022 Feb 9];n/a(n/a). Available from: https://onlinelibrary.wiley.com/doi/abs/10.1002/jmv.27604

25. Kwok SL, Cheng SM, Leung JN, Leung K, Lee C-K, Peiris JM, et al. Waning antibody levels after COVID-19 vaccination with mRNA Comirnaty and inactivated CoronaVac vaccines in blood donors, Hong Kong, April 2020 to October 2021. Eurosurveillance. 2022 Jan 13;27(2):2101197.

26. Barchuk A, Skougarevskiy D, Titaev K, Shirokov D, Raskina Y, Novkunkskaya A, et al. Seroprevalence of SARS-CoV-2 antibodies in Saint Petersburg, Russia: a population-based study. Sci Rep. 2021 Jun 21;11(1):12930.

27. Alsuwaidi AR, Al Hosani FI, Al Memari S, Narchi H, Abdel Wareth L, Kamal H, et al. Seroprevalence of COVID-19 infection in the Emirate of Abu Dhabi, United Arab Emirates: a population-based cross-sectional study. Int J Epidemiol. 2021 Aug 1;50(4):1077–90.

28. Lai C-C, Wang J-H, Hsueh P-R. Population-based seroprevalence surveys of anti-SARS-CoV-2 antibody: An up-to-date review. Int J Infect Dis. 2020 Dec 1;101:314–22.

29. Mijović B, Mašić S, Petković M, Knežević D, Aćimović J, Djaković-Dević J, et al. Seroprevalence of SARS-CoV-2 antibodies and knowledge, attitude and practice toward COVID-19 in the Republic of Srpska-Bosnia & Herzegovina: A population-based study. PLOS ONE. 2022 Jan 28;17(1):e0262738.

30. Van Pelt A, Glick HA, Yang W, Rubin D, Feldman M, Kimmel SE. Evaluation of COVID-19 Testing Strategies for Repopulating College and University Campuses: A Decision Tree Analysis. J Adolesc Health. 2021 Jan;68(1):28–34.

31. Donneau A-F, Guillaume M, Bours V, Dandoy M, Darcis G, Desmecht D, et al. University population-based prospective cohort study of SARS-CoV-2 infection and immunity (SARSSURV-ULiège): a study protocol. BMJ Open. 2022 Jan;12(1):e055721.

32. Frazier PI, Cashore JM, Duan N, Henderson SG, Janmohamed A, Liu B, et al. Modeling for COVID-19 college reopening decisions: Cornell, a case study. Proc Natl Acad Sci. 2022 Jan 11;119(2):e2112532119.

33. Cheng S-Y, Wang CJ, Shen AC-T, Chang S-C. How to Safely Reopen Colleges and Universities During COVID-19: Experiences From Taiwan. Ann Intern Med. 2020 Jul 2;M20–2927.

